# Validation and testing of a method for detection of SARS-CoV-2 RNA in healthy human stool

**DOI:** 10.1101/2020.11.09.20228601

**Authors:** Michael P. Coryell, Mikhail Iakiviak, Nicole Pereira, Pallavi P. Murugkar, Jason Rippe, David B. Williams, Jessica L. Hastie, Rosa L. Sava, Christopher Z. Lien, Tony T. Wang, William J. Muller, Michael A. Fischbach, Paul E. Carlson

**Author notes:** **Corresponding author:** Paul E. Carlson Jr. Division of Bacterial, Parasitic and Allergenic Products, Office of Vaccines Research and Review, Center for Biologics Evaluation and Research, US Food and Drug Administration,10903 New Hampshire Avenue, Silver Spring, Maryland 20993, USA.

## Abstract

**Background:** Fecal shedding of SARS-CoV-2 has raised concerns about transmission through fecal microbiota transplantation (FMT) procedures. While many tests have been authorized for diagnosis of COVID-19 using respiratory samples, no fully validated stool tests for detection of SARS-CoV-2 are currently available. We sought to adapt and validate an available test specifically for detection of SARS-CoV-2 in human stool.

**Methods:** Stool samples were spiked with inactivated SAR-CoV-2 virus for development and validation of the assay. A modified version of the CDC rRT-PCR SARS-CoV-2 test was used for detection of virus. Analytical sensitivity, assay reproducibility, and sample stability under a variety of storage conditions were assessed. We also performed the assay on stool samples collected from known COVID positive individuals.

**Findings:** The lower limit of detection (LoD) of the assay was found to be 3000 viral RNA copies per gram of original stool sample, with 100% detection across 20 replicates assessed at this concentration. Samples were relatively stable in all buffers tested at both 4°C and ambient temperature, with the exception of storage in STAR buffer at ambient temperature. Assay sensitivity was slightly diminished in low-copy-number samples after a single freeze-thaw cycle at −80°C. Thirty contrived SARS-CoV-2 samples were tested by a second laboratory and were correctly identified as positive or negative in at least one of two rounds of testing. Additionally, we detected SARS-CoV-2 RNA in the stool of known COVID-19 positive individuals using this method.

**Interpretation:** This is a sensitive, reproducible, and validated assay for detection of SARS-CoV-2 RNA in human stool with potential uses in FMT donor screening, sewage monitoring, and further research into the impact of fecal shedding on the epidemiology of this pandemic.

**Funding:** National Institute for Allergy and Infectious Diseases, NIH. Center for Biologics Evaluation and Research, FDA.

**Research in Context:** *Evidence before this study:* Since the onset of the COVID-19 pandemic, multiple studies have documented shedding of **SARS-CoV-2 RNA in feces and considered the potential for fecal-oral transmission of this virus. This potential risk led to the U**.**S. Food and Drug Administration issuing a safety alert that contained the recommendation that no stool donated after December 1, 2019 be used for manufacture of Fecal Microbiota for Transplantation (FMT) products in the United States until such a time as sufficient screening procedures could be put in place to mitigate this risk**.

*Added value of this study:* Here, we report the development and validation of an assay specifically meant for the detection of SARS-CoV-2 RNA in the stool of healthy individuals. **While studies have reported detection of viral RNA in stool previously, this is the first publication of a validated assay designed for this purpose**.

*Implications of all the available evidence:* The work presented here provides a validated SARS-CoV-2 stool **assay with potential application to FMT donor screening protocols, sewage monitoring protocols, as well as research studies assessing the role of stool shedding and transmission on the epidemiology of COVID-19**.

## Introduction

Fecal microbiota transplantation (FMT) is of interest as a treatment for numerous conditions that have been associated with dysbiosis of the gut microbiome, the most well studied of these being the treatment of *Clostridioides* (*Clostridium) difficile* infection (CDI)^1^. The screening process for stool donors includes both stool and blood testing to rule out the potential for transmission of pathogens of concern^3^. Testing recommendations for stool donors are expected to change over time, particularly as new pathogens of concern emerge. The FDA recently issued safety alerts indicating the need for increased donor screening or changes in donor testing methods used, including tests for multi-drug resistant organisms, Enteropathogenic *Escherichia coli* (EPEC) and Shiga-toxin producing *E. coli* (STEC)^4,5^.

The emergence and global spread of COVID-19 and the discovery of the SARS-CoV-2 virus in the stool of infected individuals has led to additional concerns regarding the possibility of SARS-CoV-2 transmission via FMT^6-9^. In response to early reports of stool shedding, the FDA issued a safety alert on March 23, 2020 which was later updated on April 9, 2020 to indicate that there should be no clinical use of FMT products manufactured from stool donated on or after December 1, 2019 until additional screening and testing procedures have been implemented for donor qualification programs^10,11^. This safety alert suggested possible methods for assessing donors and donor stool including routine nasal testing of donors and direct testing of donor stool for the virus. While FDA has authorized many molecular diagnostics tests for detection of SARS-CoV-2 in respiratory and oral specimen types under emergency use authorization (EUA), there are currently no EUA authorized molecular tests for stool as a specimen type. Although two publications have provided an assessment of methods for detection of SAR-CoV-2 RNA in stool^12,13^, to date, a validated stool assay with the demonstrated ability to detect SARS-CoV-2 RNA in healthy donor stool with high sensitivity has not been reported. Here, we present analytical validation of an assay for SARS-CoV-2 viral RNA detection in human stool. This testing method is a modified version of the CDC real-time reverse transcriptase PCR test that is authorized for use with respiratory samples^14^. This test does not have emergency use authorization (EUA) as a COVID-19 diagnostic, however, following individual site-specific qualification and/or validation, it may be useful as part of routine donor stool testing procedures and potentially for the release of quarantined FMT product manufactured early in the pandemic.

## Materials and Methods

### Viral reference material

Heat inactivated SARS-CoV-2 (iCOV2) was used for spike-ins. A SARS-CoV-2 isolate obtained from BEI Resources (NIAID, NIH) was propagated in Vero E6 cells (ATCC, Cat# CRL-1586) cultured in Eagle’s minimal essential medium supplemented with 10% fetal bovine serum (Invitrogen) and 1% penicillin/streptomycin and L-glutamine. Harvested cell supernatants were inactivated in a water bath at 60°C for 1 hour. Inactivated virus stocks were diluted in sterile PBS (Gibco), aliquoted, and stored at −80°C. To minimize degradation from freeze-thaw activity, fresh freezer stocks were used for each experiment.

Copy number concentrations for the virus stocks were determined by droplet digital PCR (ddPCR) using the QX-200 system (Bio-Rad), the One-Step RT-ddPCR advanced kit for probes (Bio-Rad), and 2019- nCoV CDC ddPCR Triplex probe kit (Bio-Rad) following all manufacturer protocols. RNA was isolated from freshly thawed freezer stocks using the QIAamp Viral RNA mini kit (Qiagen) following manufacturer recommendations. Absolute quantification of viral genome targets was determined using Quantasoft software V. 1.7.4 (Bio-Rad).

### Human fecal samples

Initial rRT-PCR protocol development, limit of detection experiments, and subsequent experiments were performed using human fecal material purchased from two vendors, Lee Biosolutions and OpenBiome. All fecal samples were collected by the vendors with informed consent from healthy donors between 18 and 50 years of age with no use of antibiotics for at least 30-days prior to sample donation. To ensure SARS-CoV-2 negativity, only samples collected and frozen by the vendors prior to January 2020 were purchased.

### rRT-PCR assay conditions

The protocols for isolation of viral RNA from stool samples and RNA target detection were adapted from previously published work^12^. Prior to RNA extraction, stool was diluted 1:5 (w:v) in sterile PBS, homogenized, and clarified by centrifugation at 4000 rcf for 20 minutes at 4°C. 140 μL of clarified supernatant was used as starting material for RNA extraction using the QIAamp viral RNA mini kit (Qiagen) spin-column protocol, with a final elution volume of 50 μL.

For the SARS-CoV-2 detection protocol, specimens were assayed for three unique targets in independent reactions using sequence-specific primers and fluorescent hydrolysis probes as described in the SARS-CoV-2 (2019-nCoV) CDC EUA protocol^14^. Two of the primer/probe sets, 2019_nCOV_N1 (N1) and 2019_nCOV_N2 (N2), target conserved regions within the nucleocapsid gene. A third set targeting the human RNase P gene (RP) was used as an internal amplification control. Primer/probe sets for each reaction were purchased together from IDT (2019-nCoV RUO kits).

Individual target amplifications were performed in 20 μL reactions consisting of 5 μL TaqPath™ 1-Step RT-qPCR Master Mix, CG (Life Technologies), 1·5 μL respective primer/probe mix, 8·5μL of nuclease-free water, and 5μL of extracted template RNA. The final concentrations of primers and probes were 500 nM of each primer and 125 nM of the probe. PCR reactions were performed according to the CDC EUA protocol with the exception of increasing the amplification denaturation step time from 3 to 15 seconds^14^. All PCR runs were performed on the CFX-Connect 96-well real-time PCR detection system (Bio-Rad) and real-time fluorescence data were analyzed using CFX Manager software v. 3.1 (Bio-Rad). For this protocol, amplification of either the N1 or N2 viral targets with a threshold cycle (Ct) below 40·0 was considered SARS-CoV-2 positive. Samples were classified as negative when both N1 and N2 failed to amplify with Ct < 40·0 and the RP gene target with Ct < 40·0 was successfully detected. Samples in which none of the targets amplified with Ct < 40·0 were considered invalid.

### Analytical sensitivity

An approximate range for the expected limit of detection (LoD) was determined by testing stool specimens spiked with a 10-fold dilution series of iCOV2 material (data not shown). Subsequently, negative stool slurries from two independent donors were spiked with iCOV2 reference material from a 2- fold dilution series and tested in replicates of 20 (10 per donor stool), including independent RNA extraction for each replicate. At least one un-spiked stool from each donor was tested with each batch of extractions as a negative specimen control, along with no-template controls and positive controls for each reaction. The LoD was defined as the lowest concentration at which at least 95% of samples tested positive.

Leftover spiked stool material from these dilutions was stored at −80°C and later tested to investigate the effects of freezing SARS-CoV-2 positive samples at or near the LoD. Frozen samples at 1x and 2x the empirical LoD were thawed and tested in replicates of 8 (4 per donor) to determine the impact of freeze-thaw activity on low-copy-number specimens.

### Performance evaluation

A total of 24 contrived positive specimens and 6 negative specimens were tested in duplicate rounds of testing. Positive specimens were contrived by spiking iCOV2 reference material into a negative stool matrix at approximately 1x, 2x, 5x, and 10x the LoD. Samples were prepared at the FDA and shipped over night on dry ice to Stanford University. All samples were analyzed in duplicate following the testing protocols described above. The recipient lab was blinded to the spike-in status of the specimens. Testing at Stanford was identical to the protocol used at FDA with the exception of the PCR system used (Applied Biosciences 7900 HT Fast Realtime PCR system).

Additionally, five stool specimens were obtained from four pediatric patients with recent SARS-CoV-2 positive respiratory tests (Table 3). RNA was extracted from stool at the Ann & Robert H. Lurie Children’s Hospital in Chicago, Illinois and shipped overnight on dry ice to the FDA for rRT-PCR analysis. All samples were tested in duplicate.

**Table 1.** Summary of LOD dilution testing in freshly-spiked stool specimens.

|  | Spiked stool dilutions |  |  |
| --- | --- | --- | --- |
|  | 2x LOD | 1x LOD | 0.5x LOD |
| Viral RNA copies per mL of specimen | $1.2 \times 10^3$ | $6.0 \times 10^2$ | $3.0 \times 10^2$ |
| Equivalent copies per gram stool | $6.0 \times 10^3$ | $3.0 \times 10^3$ | $1.5 \times 10^3$ |
| SARS-CoV-2 Positive* | 19/20 (95%) | 20/20 (100%) | 14/20 (70%) |
| N1 positive | 19/20 (95%) | 19/20 (95%) | 14/20 (70%) |
| Mean Ct (Std. Dev) | 33.1 (0.96) | 34.1 (1.00) | 35.1 (0.89) |
| N2 positive | 16/20 (80%) | 16/20 (80%) | 7/20 (35%) |
| Mean Ct (Std. Dev)** | 38.5 (1.49) | 38.7 (1.46) | 40.3 (1.23) |
| RP positive | 20/20 (100%) | 20/20 (100%) | 20/20 (100%) |
| Mean Ct (Std. Dev) | 31.9 (0.45) | 32.3 (0.46) | 32.2 (0.79) |

\*Positive detection of either the N1 or N2 target with Ct < 40.0
\*\*Calculations inclusive of Ct values $\geq 40.0$

**Table 2.** Summary of LOD dilution testing after single freeze-thaw.

|  | Spiked stool dilutions |  |
| --- | --- | --- |
|  | 2x LOD | 1x LOD |
| Viral RNA copies per mL of specimen | $1.2 \times 10^3$ | $6.0 \times 10^2$ |
| Equivalent copies per gram stool | $6.0 \times 10^3$ | $3.0 \times 10^3$ |
| SARS-CoV-2 Positive* | 8/8 (100%) | 6/8 (75%) |
| N1 positive | 8/8 (95%) | 4/8 (50%) |
| Mean Ct (Std. Dev) | 34.0 (1.39) | 34.1 (0.26) |
| N2 positive | 7/8 (88%) | 6/8 (75%) |
| Mean Ct (Std. Dev)** | 38.4 (1.29) | 39.6 (1.70) |
| RP positive | 8/8 (100%) | 8/8 (100%) |
| Mean Ct (Std. Dev) | 31.8 (0.68) | 32.5 (0.80) |

\*Positive detection of either the N1 or N2 target with Ct < 40.0
\*\*Calculations inclusive of Ct values $\geq 40.0$

**Table 3.** Patient information and stool testing result from clinical specimens.

| Clinical specimens tested |  |  |  |  |  |
| --- | --- | --- | --- | --- | --- |
| Specimen ID | Stool 001 | Stool 002 | Stool 003 | Stool 004** | Stool 005 |
| Patient Age | 5 years | 9 years | 5 months | 5 months | 7 months |
| Positive NP test | 3 days prior | 2 days prior* | same day | 4 days prior | 1 day prior |
| Symptoms | Asthma exacerbation | Fever, neutropenia (oncology patient) | Fever, peritonitis (with biliary atresia) | ** | Fever, diarrhea, congenital heart disease |
| Stool Test Result | Negative | Negative | SARS-CoV-2 Positive | SARS-CoV-2 Positive | SARS-CoV-2 Positive |

\*Patient had negative NP swab the day of stool collection
\*\* Same patient as sample 3

### Viral RNA stability during sample storage

The stability of SARS-CoV-2 RNA was tested in stool in different storage buffers and at different storage temperatures. Four different storage media were tested including PBS, Cary-Blair media, Stool Transport and Recovery (STAR) buffer (Roche Diagnostics), and DNA/RNA Shield (Zymo Research). Stool slurries in each buffer (1·5 grams of stool to 5 mL of buffer) were spiked with approximately 100x the LoD of inactivated SARS-CoV-2. Aliquots of each slurry were stored at either −80°C, 4°C, or ambient temperature (∼21°C). RNA extractions were performed from each sample set immediately (day 0) and on days 1, 2, 3, and 7 of storage at 4°C and ambient temperature, and on day 7 only from samples stored at −80°C. Extracted RNA was stored at −80°C until the end of the experiment, when rRT-PCR was performed in duplicate using only the N1 target. This experiment was replicated using independently collected stool samples from the same donor, for a total of two biological replicates.

### Data analysis

All reported average Ct values with accompanying standard deviations were calculated in Excel, and were inclusive of any detected Ct values, regardless of whether they met the 40 Ct cutoff for positive detection. Contrived clinical testing agreement rates and associated Clopper-Pearson confidence intervals were calculated using the Westgard QC online 2×2 contingency calculator tool following guidelines laid out in CLSI document EP12-A2^15^. Results of viral stability experiments were graphed and analyzed with mixed-effects models for repeated measures with multiple comparison testing in GraphPad Prism V. 8.4.0 (GraphPad Software). All reported p-values and confidence intervals are adjusted for multiple comparisons.

## Results

Here we sought to develop and validate an assay that could be used to reliably detect the SARS-CoV-2 RNA in stool samples. While previous work provided an assay for the detection of SARS-CoV-2 RNA in stool, that assay was developed by spiking in viral RNA, not whole virus, and it was not validated in that study^12^. Our assay uses the CDC SARS-CoV-2 primer and probe sets that have been granted Emergency Use Authorization for detection of the viral RNA in respiratory samples^14^. We have adapted these primer-probe sets for use in stool with minor modifications and validated the performance of this assay for the detection of SARS-CoV-2 RNA in human stool samples.

### Analytical performance

To validate the limit of detection of this assay, we first spiked dilutions of iCOV2 into stool and performed replicate testing of our rRT-PCR assay. We found the LoD to be 600 viral RNA copy equivalents per mL of stool matrix (Table 1). After accounting for a 1:5 stool dilution used in this protocol, this translates to 3000 copies per gram of whole stool using this extraction protocol. At viral concentrations of 1200, 600, and 300 copies per mL of stool slurry, test positivity rates were found to be 95% (19/20), 100% (20/20), and 70% (14/20), respectively (Table 1). The human RP gene was used as an amplification control in all samples to ensure that the PCR reactions were working correctly. This target was detected in all 20 replicates tested from each dilution, along with all negative specimen control extractions tests. At 1x the LoD, N1 and N2 targets were positive in 19 and 16 of the 20 replicates, respectively (Table 1). Notably, N2 amplification above baseline fluorescence was detected in all 20 samples, however only 16 met the criteria for positive detection (Ct ≤ 40·0).

The LoD in frozen samples was not consistent with that of freshly spiked samples. Frozen samples at 1x the LoD had a test positivity rate of 75% (6/8) after one freeze-thaw cycle (Table 2), suggesting that freezer storage impacts SARS-CoV-2 RNA detection in stool with low viral copy numbers. However, frozen samples at 2x the LoD tested with 100% positivity (8/8), suggesting that the effect of freezing on LoD is relatively minimal.

### Interlaboratory assessment

The reproducibility of this protocol between labs was tested using mock positive samples (iCOV2-spiked) and negative samples, which were frozen and shipped to the secondary site (Stanford University) for testing. The secondary site was blinded to the identity of these samples. Of the 30 contrived samples tested, only one was found to be invalid due to the non-amplification of all three targets. Of the 29 valid test results, 22 of 24 spiked samples tested positive, and 5 of 5 negative samples tested negative, for an overall testing agreement rate of 93·1% (CI 78·0 – 98·1). In a replicate round of testing for all samples, three of the contrived samples returned invalid results. Of the 27 valid replicate test results, 21 of 23 contrived positive samples tested positive and 4 of 4 negative samples tested negative, for an overall agreement rate of 92·6% (CI 76·6 – 97·9). All false-negative tests were in the concentration range of 1x to 2x the assay LoD, within which freezer storage can negatively impact SARS-CoV-2 detection (Table 2). Notably, there were no false-positive test results and none of the samples tested invalid or falsely negative in both rounds of replicate testing.

### Effects of storage conditions Ct signal stability

The effects of storage buffer and temperature on viral RNA stability were assessed in four stool storage buffers by spiking stool suspensions in different storage buffers at 100x the LoD. We then performed rRT-PCR on the samples and compared N1 Ct values from RNA extracted at various timepoints from each storage condition (Figure 1a). At baseline (day 0), mean Ct values were significantly higher in spiked stools suspended in Cary-Blair medium (29·4) and STAR buffer (30·4) compared with both PBS (28·8) and DNA/RNA Shield (28·5), possibly reflecting differences in the efficiency of RNA recovery following resuspension in the different buffers. Comparing samples stored at −80°C for one week with their respective baseline values, Ct values increased significantly in both PBS (29·7) and Cary-Blair (30·8), while there was no effect of freezing in the protective STAR buffer and DNA/RNA Shield (Figure 1a).

**Figure 1.**
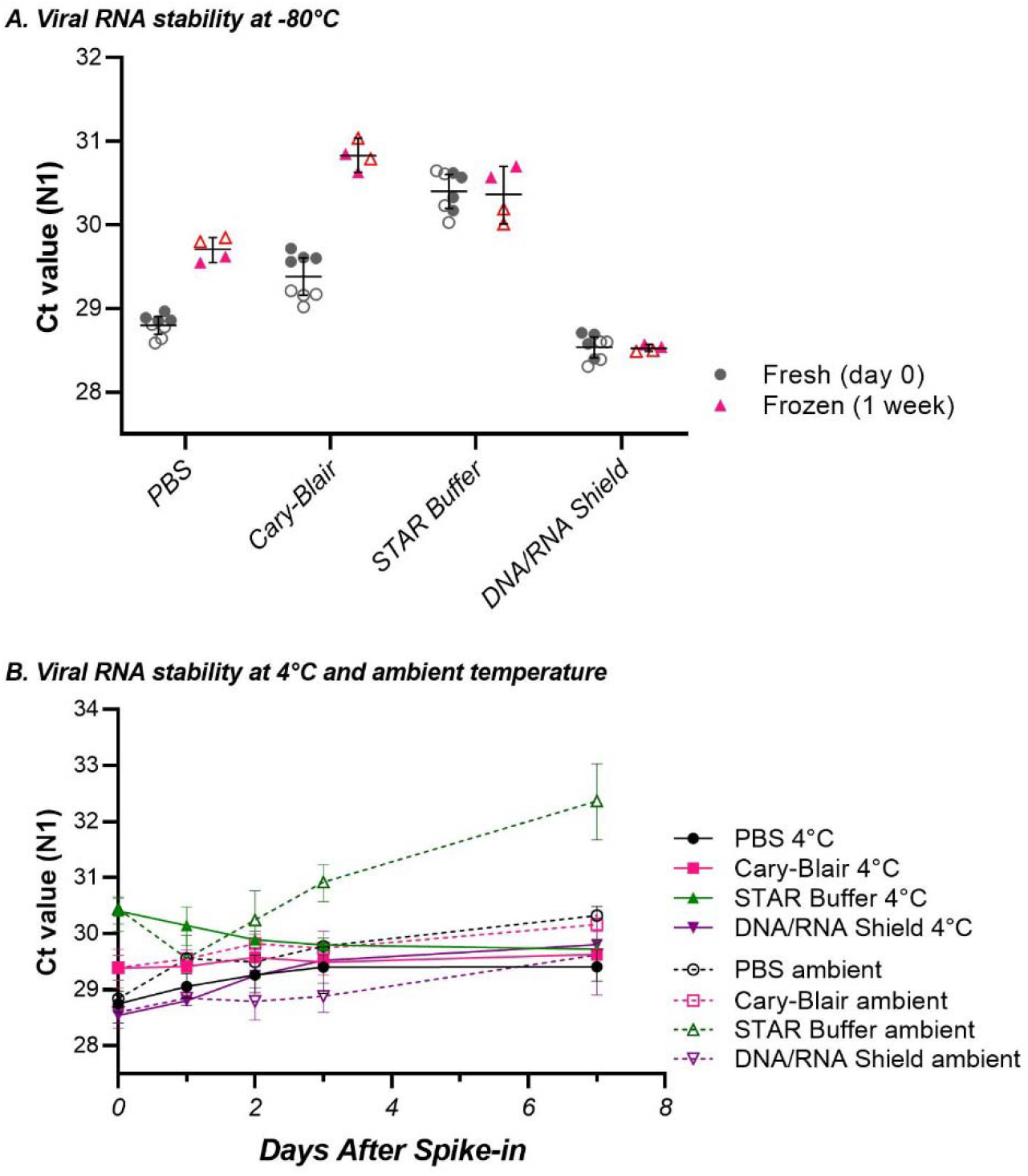
Stability of N1 Ct values in stool with different storage media and temperatures. *A*: Comparison of Ct values from freshly spiked stool (circles) with those of spiked stool frozen at −80°C for 1 week (triangles). Each symbol represents a single technical replicate, with biological replicates differentiated by solid and hollow symbol types. Bars represent simple mean and 95% confidence intervals. *B*: Ct values from stool stored at 4°C (solid) or ambient temperature (hollow). Symbols represent the mean of technical and biological replicates for a given time point and storage buffer, with 95% confidence intervals shown. ***Note: Biological replicates were assayed on separate plates but analyzed in aggregate. Baseline (day 0) values in the top and bottom panels are the same data***.

Among samples stored at 4°C for up to seven days, only those stored in DNA/RNA Shield exhibited Ct values increasing significantly above baseline, reaching significance on day 2 (p < 0·05) and a maximum Ct increase of 1·27 ± 0·29 over baseline on day 7 (Figure 1b; p < 0·001). There were non-significant increases in PBS and Cary-Blair Ct values at 4°C, each with a maximum increase at 7 days. Ct values for samples stored in STAR buffer decreased steadily compared to baseline, reaching a maximum difference of −0·65 ± 0·88 on day 7 (Figure 1b; p = 0·11). At ambient temperature storage, both PBS and STAR buffer had significant increases over baseline Ct values (Figure 1b). The maximum observed increase in STAR buffer was 1·92 ± 0·99 above baseline (p < 0·01) on day 7. In PBS, the maximum observed increase was 1·50 ± 0·04 Ct units above baseline at 7 days of storage (P ≤ 0·0001). Cary-Blair and DNA/RNA Shield samples each had non-significant increases in Ct values over baseline, each reaching maximum observed differences at 7 days (Figure 1b).

### Assessment of assay performance on clinical specimens

To definitively show that this method could detect SARS-CoV-2 RNA in the stool of infected individuals, we obtained stool samples from individuals who had previously tested positive for SARS-CoV-2 (Table 3). Five stool samples from four pediatric patients with recent COVID-19 positive respiratory swab tests were tested using our assay. Three of the five stool specimens tested positive for SARS-CoV-2 RNA, representing two of the four patients.

## Discussion

Widely documented fecal shedding of SARS-CoV-2 viral RNA, sometimes in the absence of positive respiratory testing, has led to an interest in assessing viral loads in stool for a range of purposes from diagnostics to environmental monitoring to predict outbreaks^16-19^. Additionally, the potential for transmission of SARS-CoV-2 via stool has led to concerns regarding the possible transmission as part of an FMT treatment, particularly with the possibility of asymptomatic shedding in stool^20-23^. Therefore, there is a need for SARS-CoV-2 testing to be incorporated into FMT donor screening protocols in the COVID-19 era^6,7,9^. Using accurate reference materials including human stool from healthy donors and inactivated SARS-CoV-2 virus, we have demonstrated sensitive and reproducible detection of SARS-CoV-2 RNA in stool using methods modified from the CDC RT-PCR diagnostic protocol^14^. The assay reported here has a relatively low LoD (3000 copies per gram), which is comparable to other molecular tests for enteric pathogens^24^. Further research is needed to understand the actual risk of fecal-oral transmission and the infectious dose of SARS-CoV-2 to fully evaluate the effectiveness of this assay.

The assay was able to detect SARS-CoV-2 RNA in 3 of 5 stool samples from COVID-19 patients, emphasizing the complexity between nasopharyngeal and fecal viral loads. Without access to stool from COVID-19 positive asymptomatic individuals, we performed an interlaboratory assessment using contrived samples in lieu of a clinical performance study. To prevent bias, the secondary laboratory performing the tests was blinded to the status of contrived SARS-CoV-2 positive and negative specimens.

Replicate testing results demonstrated the reproducibility of this protocol between users in different laboratories. While the blinded assessment did yield a small number of false-negative results at low viral spike-in loads, this was not unexpected based on our characterization of analytical sensitivity in frozen samples at or near the LoD. However, replicate testing improved detection sensitivity, as all samples tested correctly in at least one of two rounds of replicate testing. This provides support for incorporating replicate testing for the detection of low viral copy numbers in stool testing protocols. More work is still needed to determine the expected range of values in a study population of asymptomatic individuals infected with the SARS-CoV-2 virus. Due to a relatively small number of samples included and a lack of information regarding the expected viral load in stool of COVID-19 positive asymptomatic individuals, this evaluation was not sufficiently powered for robust estimation of clinical sensitivity or specificity.

During the early stages of the COVID-19 pandemic, high demand for testing reagents as well as challenges with cold-chain management necessitated evaluation of alternate specimen collection media and storage conditions^27^. With this in mind, we sought to assess the loss of sensitivity observed following storage in some conditions, which represents a potential limitation of this assay. To address these concerns, we evaluated the stability of virus spiked stool samples stored under a variety of conditions including different storage buffers and temperature. We assessed sample stability at a relatively high viral copy number in four storage buffers held at either 4°C or ambient temperature and found consistent detection among all media and storage temperatures. However, variation in detection signal (Ct value) suggests that both stool transport/dilution media and sample storage temperatures should be considered when developing protocols for collection, storage, and testing for SARS-CoV-2 RNA in stool. Among the media tested, DNA/RNA Shield performed best overall. It had similar baseline Ct values to PBS and provided better stability in samples stored at ambient temperature or frozen at −80°C. Notably, DNA/RNA Shield performed better at ambient temperature than at 4°C, which is consistent with the manufacturer’s recommendations for use and should be a consideration when using this product. Cary-Blair medium, a common substrate for stool transport, provided the least protection from freezing but otherwise performed similarly to PBS at 4°C. The results following freezing of spiked stool samples did show potential negative effects on sample stability. Notably, freezer storage of spiked specimens reduced detection in low-copy-number samples. This was also apparent in our reproducibility evaluation in which some low-copy-number frozen specimens returned false-negative results. Since we were unable to evaluate viral RNA stability in stool from known positive donors, it is not known whether freezing of raw stool samples would have a similar effect on SARS-CoV-2 RNA detection.

While the potential transmission of SARS-CoV-2 via the fecal-oral route and in FMT remains a topic of some debate, there have been reports showing clear evidence of gastrointestinal infection, including isolation of infectious virus from stool^17,20^. Here, we present a technical validation of methods developed for screening stool for SARS-CoV-2 viral RNA. In contrast to testing protocols published by the CDC and others, we considered detection of either N1 or N2 viral target sufficient for a SARS-CoV-2 RNA positive test determination, regardless of RP amplification. Some of our results were consistent with previous reports that the N1 primer/probe set is more sensitive than N2^28^. However, our data include several valid results which were N2 positive and N1 negative, supporting the continued use of N2 in SARS-CoV-2 testing. This study did not investigate the use of alternative SARS-CoV-2 RT-PCR primer/probe sets, nor did we explore alternative testing technologies like loop-mediated isothermal amplification (a.k.a. LAMP) and CRISPR-guided detection methods. Taken together, the data presented here demonstrate a validated stool detection method for SARS-CoV-2 RNA that has the potential for use in a variety of applications including FMT donor screening, sewage monitoring, and clinical research.

## Data Availability

All data generated for this study are presented in the manuscript.

## Contributors

MC, JH, WM, and PC designed the study. MC performed assay development, validation and implementation. MC, MF, and PC analyzed the data. MI, NP, PM, and MF performed secondary site implementation and testing. CL and TW generated inactivated virus used in the studies. JR, DW, and WM acquired and processed clinical samples. MC, JH, RS, MF, and WM assisted in manuscript preparation. All authors reviewed the final manuscript.

## Acknowledgements

This work was supported by an Inter-Agency Agreement to PC between the National Institutes for Health and Food and Drug Administration (AAI17016-001-00001) and the Intramural Research Program of the Center for Biologics Evaluation and Research, Food and Drug Administration. This project was supported in part by an appointment to the Research Fellowship Program at the OVRR/CBER, U.S. Food and Drug Administration, administered by the Oak Ridge Institute for Science and Education through an interagency agreement between the U.S. Department of Energy and FDA.

## References

1. Choi HH, Cho YS. Fecal Microbiota Transplantation: Current Applications, Effectiveness, and Future Perspectives. Clin Endosc 2016; 49(3): 257–65.

2. US Food and Drug Administration. Enforcement Policy Regarding Investigational New Drug Requirements for Use of Fecal Microbiota for Transplantation to Treat Clostridium difficile Infection Not Responsive to Standard Therapies. https://www.fda.gov/media/86440/download; 2013.

3. Carlson PE, Jr. Regulatory Considerations for Fecal Microbiota Transplantation Products. Cell Host Microbe 2020; 27(2): 173–5.

4. US Food and Drug Administration. Information Pertaining to Additional Safety Protections Regarding Use of Fecal Microbiota for Transplantation -- Testing of Stool Donors for Enteropathogenic Escherichia coli and Shigatoxin-Producing Escherichia coli. https://www.fda.gov/vaccines-blood-biologics/safety-availability-biologics/information-pertaining-additional-safety-protections-regarding-use-fecal-microbiota-transplantation-0 ; 2020.

5. US Food and Drug Administration. Safety Alert Regarding Use of Fecal Microbiota for Transplantation and Risk of Serious Adverse Events Likely Due to Transmission of Pathogenic Organisms. https://www.fda.gov/vaccines-blood-biologics/safety-availability-biologics/safety-alert-regarding-use-fecal-microbiota-transplantation-and-risk-serious-adverse-events-likely ; 2020.

6. Ianiro G, Mullish BH, Kelly CR, et al. Reorganisation of faecal microbiota transplant services during the COVID-19 pandemic. Gut 2020; 69(9): 1555–63.

7. Chiu CH, Tsai MC, Cheng HT, L. PH, Kuo CJ, Chiu CT. Fecal microbiota transplantation and donor screening for Clostridioides difficile infection during COVID-19 pandemic. J Formos Med Assoc 2020.

8. Ianiro G, Mullish BH, Kelly CR, et al. Screening of faecal microbiota transplant donors during the COVID-19 outbreak: suggestions for urgent updates from an international expert panel. Lancet Gastroenterol Hepatol 2020; 5(5): 430–2.

9. Green CA, Quraishi MN, Shabir S, et al. Screening faecal microbiota transplant donors for SARS-CoV-2 by molecular testing of stool is the safest way forward. Lancet Gastroenterol Hepatol 2020; 5(6): 531.

10. US Food and Drug Administration. Safety Alert Regarding Use of Fecal Microbiota for Transplantation and Additional Safety Protections Pertaining to SARS-CoV-2 and COVID-19. https://www.fda.gov/vaccines-blood-biologics/safety-availability-biologics/safety-alert-regarding-use-fecal-microbiota-transplantation-and-additional-safety-protections ; 2020.

11. US Food and Drug Administration. Information Pertaining to Additional Safety Protections Regarding Use of Fecal Microbiota for Transplantation - Screening Donors for COVID-19 and Exposure to SARS-CoV-2 and Testing for SARS-CoV-2. https://www.fda.gov/vaccines-blood-biologics/safety-availability-biologics/information-pertaining-additional-safety-protections-regarding-use-fecal-microbiota-transplantation-1 ; 2020.

12. Ng SC, Chan FKL, Chan PKS. Screening FMT donors during the COVID-19 pandemic: a protocol for stool SARS-CoV-2 viral quantification. Lancet Gastroenterol Hepatol 2020; 5(7): 642–3.

13. Perchetti GA, Nalla AK, Huang ML, et al. Validation of SARS-CoV-2 detection across multiple specimen types. J Clin Virol 2020; 128: 104438.

14. Centers for Disease Control and Prevention Division of Viral Diseases. CDC 2019-Novel Coronavirus (2019-nCoV) Real-Time RT-PCR Diagnostic Panel https://www.cdc.gov/coronavirus/2019-ncov/lab/rt-pcr-panel-primer-probes.html; 2020.

15. Garrett PE LF, Meier KL. User protocol for evaluation of qualitative test performance: approved guideline.: Clinical and Laboratory Standards Institute,; 2008.

16. Randazzo W, Truchado P, Cuevas-Ferrando E, Simon P, Allende A, Sanchez G. SARS-CoV-2 RNA in wastewater anticipated COVID-19 occurrence in a low prevalence area. Water Res 2020; 181: 115942.

17. Xiao F, Sun J, Xu Y, et al. Infectious SARS-CoV-2 in Feces of Patient with Severe COVID-19. Emerg Infect Dis 2020; 26(8): 1920–2.

18. Cheung KS, Hung IFN, Chan PPY, et al. Gastrointestinal Manifestations of SARS-CoV-2 Infection and Virus Load in Fecal Samples From a Hong Kong Cohort: Systematic Review and Meta-analysis. Gastroenterology 2020; 159(1): 81–95.

19. Yu F, Yan L, Wang N, et al. Quantitative Detection and Viral Load Analysis of SARS-CoV-2 in Infected Patients. Clin Infect Dis 2020; 71(15): 793–8.

20. Xiao F, Tang M, Zheng X, Liu Y, Li X, Shan H. Evidence for Gastrointestinal Infection of SARS-CoV-2. Gastroenterology 2020; 158(6): 1831–3 e3.

21. van Doorn AS, Meijer B, Frampton CMA, Barclay ML, de Boer NKH. Systematic review with meta-analysis: SARS-CoV-2 stool testing and the potential for faecal-oral transmission. Aliment Pharmacol Ther 2020.

22. Teixeira SC. Mild and asymptomatic cases of COVID-19 are potential threat for faecal-oral transmission. Braz J Infect Dis 2020; 24(4): 368.

23. Hindson J. COVID-19: faecal-oral transmission? Nat Rev Gastroenterol Hepatol 2020; 17(5): 259.

24. Harrington SM, Buchan BW, Doern C, et al. Multicenter evaluation of the BD max enteric bacterial panel PCR assay for rapid detection of Salmonella spp., Shigella spp., Campylobacter spp. (C. jejuni and C. coli), and Shiga toxin 1 and 2 genes. J Clin Microbiol 2015; 53(5): 1639–47.

25. Rose C, Parker A, Jefferson B, Cartmell E. The Characterization of Feces and Urine: A Review of the Literature to Inform Advanced Treatment Technology. Crit Rev Environ Sci Technol 2015; 45(17): 1827–79.

26. Biswas B. Clinical Performance Evaluation of Molecular Diagnostic Tests. J Mol Diagn 2016; 18(6): 803–12.

27. Rogers AA, Baumann RE, Borillo GA, et al. Evaluation of Transport Media and Specimen Transport Conditions for the Detection of SARS-CoV-2 by Use of Real-Time Reverse Transcription-PCR. J Clin Microbiol 2020; 58(8).

28. Vogels CBF, Brito AF, Wyllie AL, et al. Analytical sensitivity and efficiency comparisons of SARS-CoV-2 RT-qPCR primer-probe sets. Nat Microbiol 2020; 5(10): 1299–305.

